# Rapid implementation of SARS-CoV-2 emergency use authorization RT-PCR testing and experience at an academic medical institution

**DOI:** 10.1101/2020.06.05.20109637

**Authors:** P. Velu, A. Craney, P. Ruggiero, J. Sipley, L. Cong, Erika M. Hissong, M. Loda, L. F. Westblade, M. Cushing, H. Rennert

## Abstract

An epidemic caused by an outbreak of severe acute respiratory syndrome coronavirus 2 (SARS-CoV-2) in China in December 2019 has since rapidly spread internationally, requiring urgent response from the clinical diagnostics community. We present a detailed overview of the clinical validation and implementation of the first laboratory-developed real-time reverse-transcription-PCR (rRT-PCR) test offered in the NewYork-Presbyterian Hospital system following the emergency use authority (EUA) guidance issued by the US Food and Drug Administration. Validation was performed on nasopharyngeal and sputum specimens (n=124) using newly designed dual-target rRT-PCR (altona RealStar® SARS-CoV-2 Reagent) for detecting of SARS-CoV-2 in upper respiratory and lower respiratory tract specimens, including bronchoalveolar lavage and tracheal aspirates. Accuracy testing demonstrated excellent assay agreement between expected and observed values. The limit of detection (LOD) was 2.7 and 23.0 gene copies/reaction for nasopharyngeal and sputum specimens, respectively. Retrospective analysis of 1,694 tests from 1,571 patients revealed increased positivity in older patients and males compared to females, and an increasing positivity rate from approximately 20% at the start of testing to 50% at the end of testing three weeks later. Our findings demonstrate that the assay accurately and sensitively identifies SARS-CoV-2 in multiple specimen types in the clinical setting and summarizes clinical data from early in the epidemic in New York City.

## INTRODUCTION

The novel coronavirus SARS CoV-2 is a member of the *Betacoronavirus* genera in the subfamily *Coronavirinae*, which is known to cause respiratory illness and gastroenteritis in humans and other mammals [1, 2]. Two other *Betacoronavirus* that have met with global attention are SARS-CoV (2002) and MERS-CoV (2012). An outbreak of respiratory disease caused by SARS-CoV-2, first detected in Wuhan, China at the end of December 2019, rapidly spread to other countries, including the United States [3, 4], resulting in New York City in particular becoming an epicenter of the pandemic [5]. Given the devastating impact on the healthcare system and the need for accurate and quick diagnosis of SARS-CoV-2 infection the United States Food and Drug Administration (FDA) has established a rapid pathway for using laboratory-developed tests (LDTs) that was outlined in a guidance document published on February 29, 2020 [6]. According to this guidance SARS-CoV-2 testing may be performed by CLIA-certified high-complexity molecular laboratories under Emergency Use Authorization (EUA) according to a set of recommendations regarding the minimum validation required for ensuring the analytical and clinical validity of the test. Details of the test and validation must be submitted by the laboratory to the FDA through an EUA application within 15 days of initiating testing, after which testing may continue provisionally until a decision by the FDA is rendered.

The Center for Disease Control (CDC) and the New York State Department of Health (NYS DOH) had designed and manufactured new test kits for SARS-CoV-2. However, very few laboratories were able to get access to these reagents, and their use has been limited by the need for specific instruments, which were not available in our institution. Additionally, limited access to SARS-CoV-2 RNA reference control material presented a significant hurdle to the validation process. The FDA EUA announcement allowed laboratories to procure SARS-CoV-2 RNA from the World Reference Center for Emerging Viruses and Arboviruses (WRCEVA) or the National Institutes of Health (NIH) Biodefense and Emerging Infections Research Resources Repository (BEI).

The scale of demand for diagnostic testing and the shortage of supplies led to the need for a high throughput diagnostic test that could be readily implemented in a variety of laboratories. Here we describe how in less than five days, our institution validated and submitted for FDA EUA approval the research use only (RUO) RealStar® SARS-CoV-2 Reagent Kit 1.0 (Altona Diagnostics) test. Comparable validation studies that can take months were completed within a week. We also detail workflow considerations and results from three weeks of testing from March 11, 2020, through March 31, 2020, during the early days of the COVID-19 outbreak in New York City.

## MATERIALS AND METHODS

### SARS-CoV-2 RNA control material

We obtained SARS-CoV-2 RNA reference material from WRCEVA (University of Texas Medical Branch, Galveston, TX, Strain USA_WA1/2020, Lot TVP 23156, RNA preparation date 2/21/2020) for use in clinical evaluation and limit of detection (LOD) studies. We used this RNA reference material to perform LOD dilution series experiments and to create contrived positive samples for accuracy studies by spiking it into pooled leftover negative patient specimens.

### Validation samples and clinical cohort

An in-house validation panel consisting of a total of 124 contrived samples and patient specimens, including NP (64) and sputum (60) specimen types, was used for the validation. Samples were obtained from individuals suspected of respiratory tract infections. All NP samples had been clinically tested for the presence of twenty-one common respiratory viruses using our institution’s respiratory virus panel, the commercially available BioFire FilmArray® Respiratory Pathogen 2 (RP2) panel (BioFire Diagnostics, LLC, Salt Lake City, USA). Reactive clinical samples consisted of four patient specimens confirmed to have SARS-CoV-2 by the New York City-Department of Health and Mental Hygiene (NYC-DOH) using the NYS-DOH SARS-CoV-2 EUA assay and samples contrived by spiking WRCEVA RNA material into pooled leftover negative clinical specimens.

Additionally, we performed a retrospective analysis of patient characteristics on 1,694 consecutive upper respiratory tract (URT) specimens tested on the assay obtained from 1,571 patients with high suspicion for COVID-19 that were treated at NewYork Presbyterian Hospital (NYPH) campuses from March 11 to March 31, 2020. The IRB Committee at Weill Cornel Medicine (WCM) approved this study.

### Real time reverse-transcription PCR testing

Automated extraction of total nucleic acid (TNA) was performed on 200 μl of NP swab viral transport media (VTM) following an off-board lysis viral inactivation step, using the QIAsymphony DSP Virus/Pathogen Mini Kit coupled on the QIAsymphony SP (Qiagen, Germantown, MD), with a resulting eluate volume of 60 μL. The viral inactivation step was performed in a class 2 biosafety cabinet using personal protective equipment following our hospital biosafety policies. For sputum, 100 μL of specimen was first treated with 0.3% dithiothreitol (DTT) solution (1:1 ratio) and incubated at 37° C for 30 minutes to reduce viscosity. One-step reverse transcription to cDNA and rRT-PCR of viral targets (Envalope and Spike genes) and internal control (IC) was performed using 10 μL TNA eluate and the RealStar® SARS-CoV-2 real-time RT-PCR Kit 1.0 (altona Diagnostics Gmbh, Hamburg, Germany) on the Rotor-Gene Q Thermocyler (Qiagen) for a total volume of 30 μL per reaction. PCR amplification and detection were performed using multi-color fluorescent dye-labeled probes for the identification and differentiation of B-betacoronavirus (B-βCoV) and SARS-CoV-2 specific RNA and the detection of the IC within one reaction, allowing for higher throughput testing compared to the CDC assay. Samples in which both the E gene target (all B-βCoV) and the S gene target (SARS-CoV-2 specific), or the S gene target only were detected within the first 40 cycles of amplification were considered “Detected”. Samples with cycle threshold (Ct) values ≥ 40.0 were considered negative. Each run contained an external positive control (1:10^3^ WRCEVA RNA dilution), positive kit control (synthetic B-βCoV and SARS-CoV-2 RNA), SARS-CoV-2 NP negative control, and a non-template control (NTC).

### Assay Performance characteristics

The FDA EUA mechanism specified four distinct performance characteristics consisting of limit of detection (LOD), inclusivity (analytical sensitivity) cross-reactivity (analytical specificity), and clinical evaluation (accuracy) studies. For the LOD studies, SARS-CoV-2 inactivated virus or RNA spiked into artificial or real clinical matrix was acceptable, as long as the matrix was from the most difficult specimen type accepted for testing on the clinical assay (in decreasing order of difficulty: sputum, other lower respiratory tract specimens, and NP or oropharyngeal (OP) swabs collected and transported in viral transport media). NP and sputum samples were tested on the altona RealStar® rRT-PCR assay to ensure the absence of SARS-CoV-2 and pooled for use as a matrix for spiking in RNA for LOD studies and accuracy studies. Six ten-fold serial dilutions (1 × 10^1^ to 1 × 10^7^) were performed with three replicates at each concentration by spiking WRCEVA RNA reference material (60,000 pfu/μL stock WRCEVA RNA reference material; ~6:10^7^ genomic copies/μL) into NP and sputum eluates obtained from pooled-negative patient NP or sputum specimens. The dilutions at the LOD were performed by spiking 1 μl of 1:10^5^ dilution to 60 μL eluate, yielding a concentration of 0.01 pfu/uL=10pfu/mL (~10,000copies/mL). Assay performance at the determined LOD was confirmed with at least 20 additional replicates for each type of sample (sputum and NP).

For the accuracy studies, a total of 64 positive (34 NP and 30 sputum specimens, respectively) specimens and 60 negative (30 NP swabs and 30 sputum) specimens that tested negative for SARS-CoV-2 were used. Positive specimens were either contrived positive samples generated by spiking WRCEVA RNA reference material into pooled leftover negative NP or sputum specimens or real patient specimens, as described above. Twenty of the contrived clinical specimens were spiked at a concentration of 1x-2x LoD, with the remainder of samples spanning the assay testing range. FDA defines the acceptance criteria for the performance as 95% agreement at 1x-2x LOD, and 100% agreement at all other concentrations and negative specimens [6].

Inclusivity and cross-reactivity studies used a combination of *in silico* and *in vitro* approaches. As the primer and probe sequences were proprietary to the kit manufacturer, we included the results of their *in silico* analysis in our EUA application. Additional studies to determine cross-reactivity were performed in our laboratory by testing 10 NP samples that were positive by the RP2 for the four human coronaviruses NL63 (n=2), 229E (n=2), OC43 (n=4), or HKU1 (n=2), defined as high priority pathogens from the same genetic family by the FDA.

### Data analysis and statistical methods

Data analyses, including statistics and plot generation, were performed using R programming language v 3.6.0 [7]. LOD was determined through a probit regression model using the glm function following CLSI EP17A2E Guidance with Application to Quantitative Molecular Measurement Procedures [8].

## RESULTS

### Validation of Assay

#### a. Limit of detection

Dilution series studies on pooled negative NP specimens spiked with WRCEVA RNA reference material, with three replicates across a viral range of 1 gene copy/reaction to 1, 000,000 gene copies/reaction (1 to 6 log_10_), demonstrated an accurate and linear response across five logs of detection for NP and four logs of detection for sputum (**Table 1, Figure 1**). Probit analysis was applied to the NP data after an additional five replicates of testing were performed at 0.8, 0.6, 0.5, 0.4, and 0.2 gene copies/reaction, and narrowed the LOD to 2.7 gene copies/reaction at 95% detection rate (**Figure 2**). A similar LOD series and probit analysis was performed on sputum at 80, 60, 50, 40, and 20 gene copies/reaction, and resulted in a lower sensitivity with a LOD of 23.0 gene copies/reaction at 95% detection rate in sputum compared to NP specimens. For both NP and sputum specimens, 20/20 and 23/23 additional replicates tested at their LODs, respectively, resulted as positive.

**Table 1.** Limit of detection studies were performed for NP VTM (NP) and sputum (SPU) specimen types with three replicates at each dilution. An additional twenty (NP) and twenty-three (SPU) specimens were tested at the estimated LOD of 10 gene copies/reaction and 100 gene copies/reaction, respectively.

| Dilution | Gene copies per reaction | Number run |  | Number detected |  | Percent |  | Cy5 (S gene) Mean Ct |  | FAM™ (E gene) Mean Ct |  | JOE™ (I.C.) Mean Ct |  |
| --- | --- | --- | --- | --- | --- | --- | --- | --- | --- | --- | --- | --- | --- |
|  |  | NP | SPU | NP | SPU | NP | SPU | NP | SPU | NP | SPU | NP | SPU |
| 1:10 | 10<sup>6</sup> | 3 | 3 | 3 | 3 | 100% | 100% | 14.6 | 13.1 | 15.2 | 14.5 | 30.6 | 33.0 |
| 1:10<sup>2</sup> | 10<sup>5</sup> | 3 | 3 | 3 | 3 | 100% | 100% | 18.0 | 16.5 | 18.7 | 17.9 | 29.0 | 30.4 |
| 1:10<sup>3</sup> | 10<sup>4</sup> | 3 | 3 | 3 | 3 | 100% | 100% | 20.9 | 19.9 | 21.6 | 21.4 | 28.3 | 30.8 |
| 1:10<sup>4</sup> | 10<sup>3</sup> | 3 | 3 | 3 | 3 | 100% | 100% | 24.6 | 23.8 | 25.2 | 25.3 | 29.4 | 29.7 |
| 1:10<sup>5</sup> | 10<sup>2</sup> | 3 | 26 | 3 | 26 | 100% | 100% | 27.9 | 31.1 | 28.4 | 31.0 | 30.2 | 29.8 |
| 1:10<sup>6</sup> | 10 | 23 | 3 | 23 | 0 | 100% | 0% | 32.3 | ND | 32..0 | ND | 30.8 | 29.7 |
| 1:10<sup>7</sup> | 1 | 3 | 3 | 0 | 0 | 0% | 0% | ND | ND | ND | ND | 29.8 | 29.8 |

**Figure 1.**
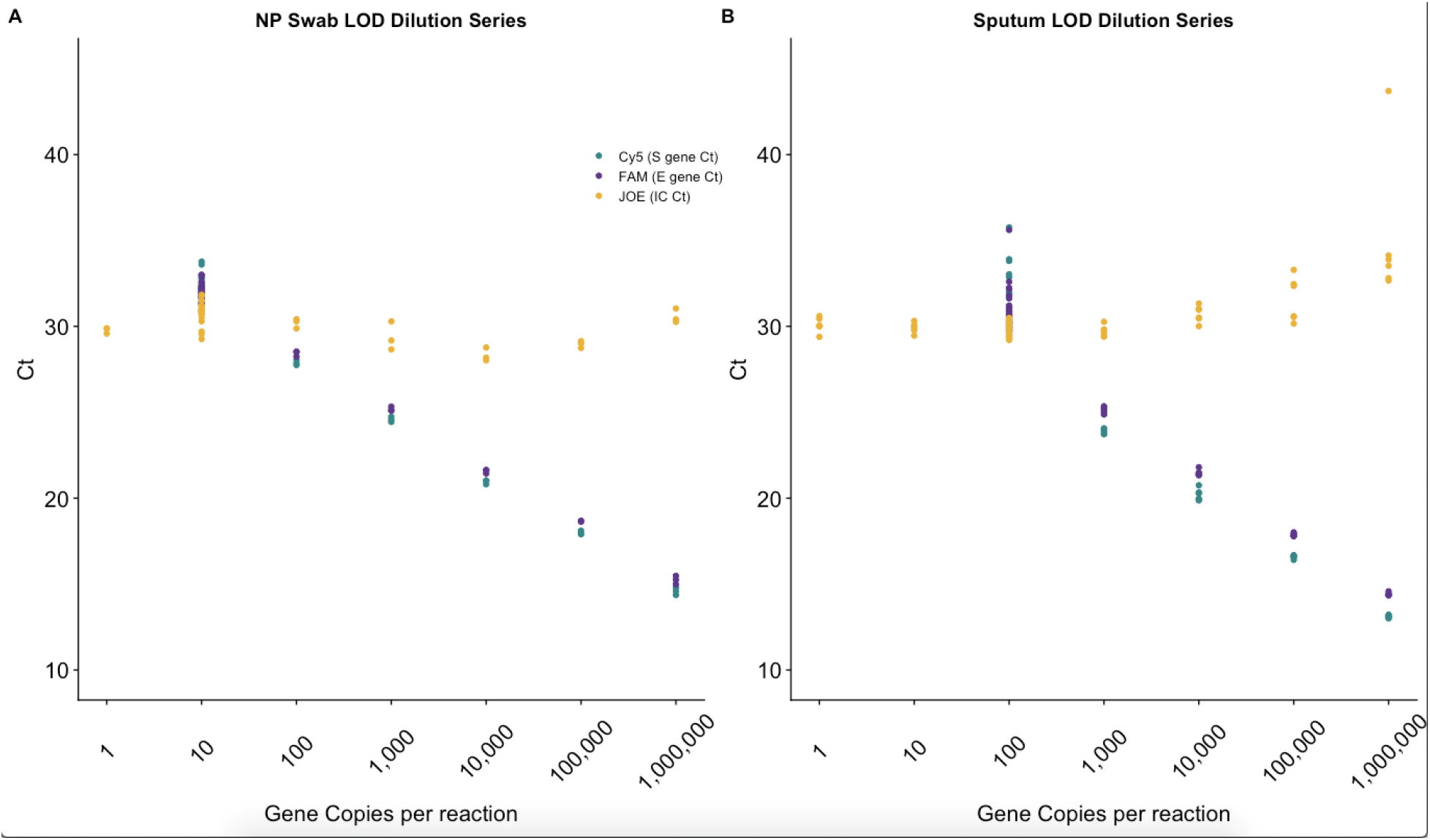
Limit of detection (LOD) studies. Ct values for the LOD serial dilution study using WRCEVA RNA reference material spiked in pooled negative **(A)** nasopharyngeal (NP) specimen eluate and **(B)** sputum specimen eluate. Six ten-fold dilutions were performed starting at 1,000,000 gene copies/reaction and ending at 1 gene copy/reaction. The apparent LOD was between 1 and 10 gene copies/reaction for NP specimens and between 10 and 100 gene copies/reaction for sputum specimens.

**Figure 2.**
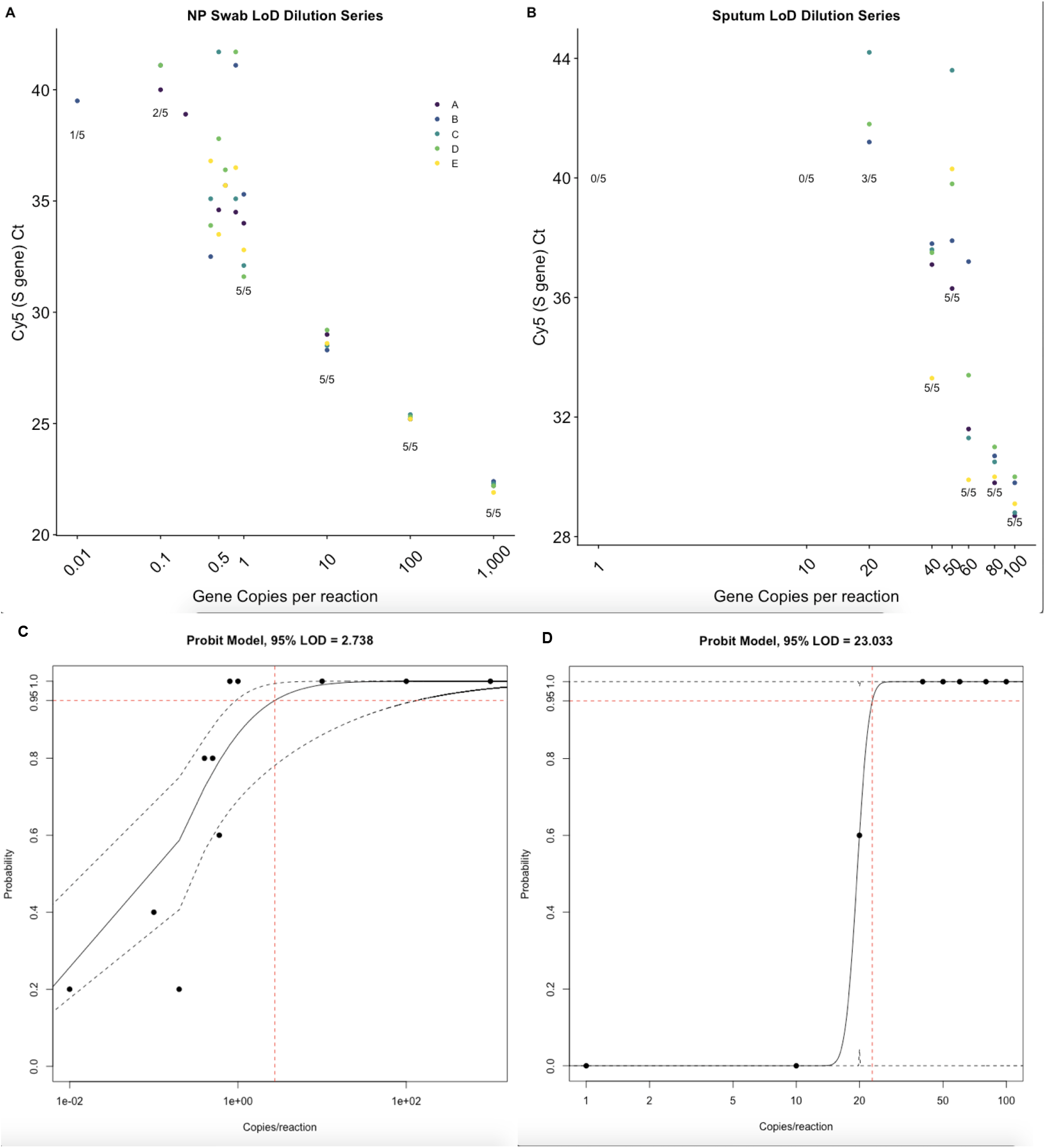
LOD of NP and sputum by probit analysis. Additional serial dilution studies were performed using WRCEVA RNA reference material spiked in pooled negative **(A)** NP specimen eluate and **(B)** sputum specimen eluate to determine the LOD. Five replicates (A, B, C, D, E) of six ten-fold dilutions were performed starting at 1,000 gene copies/reaction and ending at 0.1 gene copies/reaction for NP and five replicates of three ten-fold dilutions were performed starting at 100 gene copies/reaction and ending at 1 gene copies/reaction for sputum. An additional five replicates were performed at 0.8, 0.6, 0.5, 0.4, and 0.2 gene copies/reaction for NP and 80, 60, 50, 40, and 20 gene copies/reaction for sputum. Probit analysis showed LOD to be **(C)** 2.7 gene copies/reaction for NP and **(D)** 23.0 gene copies/reaction for sputum.

#### b. Inclusivity and Specificity

The *in silico* analysis for inclusivity that was performed by the manufacturer of the kit found 100% homology of the E gene and S gene forward and reverse primers and probes with 563 whole-genome sequences of SARS-CoV-2 published in GISAID and NCBI as of 3/16/2020 [9]. The *in silico* analysis for cross-reactivity that was performed by the manufacturer found that the S gene and E gene forward and reverse primers and probes had less than 80% homology with the vast majority of 40 different pathogens (125 strains total) tested (see Supplementary Material). In cases with greater than 80% homology, cross-reactivity was not a concern as only the forward or reverse primer, but never both primers were affected, thus rendering amplification impossible. Additionally, all ten samples that had human coronaviruses NL63, 229E, OC43, or HKU1 detected by RP2 panel tested negative for SARS-CoV-2 on the RealStar® rRT-PCR assay.

#### c. Clinical evaluation-Accuracy

All 30 NP specimens that tested negative on the RP2 panel also tested negative on the SARS-CoV-2 rRT-PCR assay (**Table 2**). Leftover VTM from these negative specimens was pooled to create a sample matrix for the LOD and contrived positive sample studies. Clinical evaluation studies resulted in the detection of SARS-CoV-2 in all specimens contrived by spiking WRCEVA RNA reference material (n=20) into pooled SARS-CoV-2 negative NP VTM or sputum and all four positive patient samples tested by NYC-DOH (**Table 2** and **Supplementary Table 1**). The high-positive patient sample run at successive dilutions (n=10) remained positive throughout the range of concentrations (1:2 to 1:1,024; Ct range, 22-31) (**Figure 1**). Similar clinical evaluation studies were performed for sputum specimens (**Table 2** and **Supplementary Table 2**), also with 100% concordance. Validation studies performed on bronchoalveolar lavage (BAL) and tracheal aspirate specimens and additional sample collection systems (**Table 3** and **Supplementary Material including Tables 3 and 4**) showed 100% accuracy.

**Table 2.** Accuracy studies for NP and sputum. Clinical evaluation of the RealStar® SARS-CoV-2 rRT-PCR assay using automated QIAsymphony total nucleic acid (TNA) extraction followed by rRT-PCR targeting the E and S coronavirus genes on an in-house validation panel consisting of patient and contrived samples for NP (124) and sputum (62).

| Validation sample | altoma SARS-CoV2 rRT-PCR |  |  |  |
| --- | --- | --- | --- | --- |
|  | NP-VTM |  | Sputum |  |
|  | Positive | Negative | Positive | Negative |
| *Positive (contrived) | 30 | 0 | 30 | 0 |
| Positive (patient) | 4 | 0 | 1 | 0 |
| Negative (patient) | 0 | 30 | 0 | 30 |
| Total | 64 | 60 | 31 | 31 |

\*SARS-CoV-2 contrived positive samples generated with RNA control spiked into pooled negative eluate or dilutions of a high positive clinical sample.

**Table 3.** Summary table of accuracy studies for all specimen types. Mean and range of Ct values are shown for positive samples. The number of positive (either contrived through spiking RNA into a negative matrix or actual patient samples) and negative specimens are also noted along with the percent of specimens that were correctly classified as positive or negative.

| **Specimen Type** | **Cy5 (S gene)**<br>**Mean Ct** | **FAM™ (E gene)**<br>**Mean Ct** | **JOE™ (I.C.)**<br>**Mean Ct** | **Number tested** | **Correctly classified** |
| --- | --- | --- | --- | --- | --- |
| NP swab | 28.8 (20.6-37.2) | 28.8 (20.8-37.7) | 29.6 (28.6-30.8) | 34+/30- | 100% |
| Sputum | 24.1 (16.2-30.0) | 24.2 (17.0-29.2) | 30.2 (29.4-32.3) | 31+/30- | 100% |
| BAL | 23.4 (16.9-29.3) | 23.5 (17.1-29.4) | 30.4 (28.6-32.4) | 20+/20- | 100% |
| Trach asp | 22.1 (10.0-43.0) | 19.7 (10.1-32.1) | 34.1 (29.7-42.4) | 5+/5- | 100% |

**Table 4.** Summary table of patient characteristics. For race, “Declined” and “Other” categories were not used when performing the Chi-squared test for significance. An additional 63 tests were performed at low numbers at several other locations; these were not included in the table.

| Feature | All Patients<br>n=1579 | Positive<br>38% | Negative<br>62% | p-value |
| --- | --- | --- | --- | --- |
| Age<br>(years) | Overall: 53.4 (0.1-120.3) | Overall: 57.5 (1.3-120.3) | Overall: 51.1 (0.1-97.5) | 0.0005* |
|  | 0-18 80 | 0-18 7% | 0-18 93% |  |
|  | 19-35 295 | 19-35 30% | 19-35 70% |  |
|  | 36-55 420 | 36-55 40% | 36-55 60% |  |
|  | 56-85 656 | 56-85 47% | 56-85 53% |  |
|  | >85 128 | >85 30% | >85 70% |  |
| Gender | F 784 | F 31% | F 69% | 0.005* |
|  | M 778 | M 46% | M 54% |  |
|  | Unsp. 3 | Unsp. 33% | Unsp. 66% |  |
| Race | Asian 101 | Asian 28% | Asian 72% | 0.385 |
|  | Black 171 | Black 36% | Black 64% |  |
|  | Declined 173 | Declined 40% | Declined 60% |  |
|  | Other 210 | Other 45% | Other 55% |  |
|  | White 488 | White 32% | White 68% |  |
| Location | Emergency 911 | Emergency 50% | Emergency 50% | 0.0005* |
|  | Inpatient 492 | Inpatient 18% | Inpatient 82% |  |
|  | Outpatient 113 | Outpatient 35% | Outpatient 65% |  |

### Clinical cohort characterization

The Altona rRT-PCR SARS-CoV-2 test was used to test NP and OP swabs from March 11, 2020, to March 30, 2020. Starting March 30, 2020, our institution deployed the higher throughput Roche cobas 6800 SARS-CoV-2 rRT-PCR test (Roche Molecular Systems, Inc; Branchburg, NJ) [10] to meet increasing specimen volumes. During the initial phase with only Altona testing, 1,694 tests were performed on 1,372 NP (40% positive), 57 OP (19% positive), and 311 NP/OP (25% positive) swab specimens from 1,571 patients. Ct values were not significantly different for the E gene, S gene, and IC targets between positive NP, OP, or NP/OP samples (**Supplementary Figure 1**). The number of tests with indeterminate or B-βCoV results were 5 and 4, respectively, all in NP swab samples. The mean Ct for E gene, S gene, and IC targets in positive samples were 23.0 (11.1-40.7), 22.5 (10.3-40.6), and 29.6 (27.0-38.4), respectively (**Figure 3**). Using Ct value of the S target gene as a surrogate for viral burden, the upper respiratory tract specimens could be classified into three groups: high (Ct <20; n=222, 34%), medium (Ct 20 - 30, n =335, 52%), and low (Ct > 30, n = 89, 13.8%). Over three weeks of testing, more than 75% of positive samples could be classified as having medium to high viral burden.

**Figure 3.**
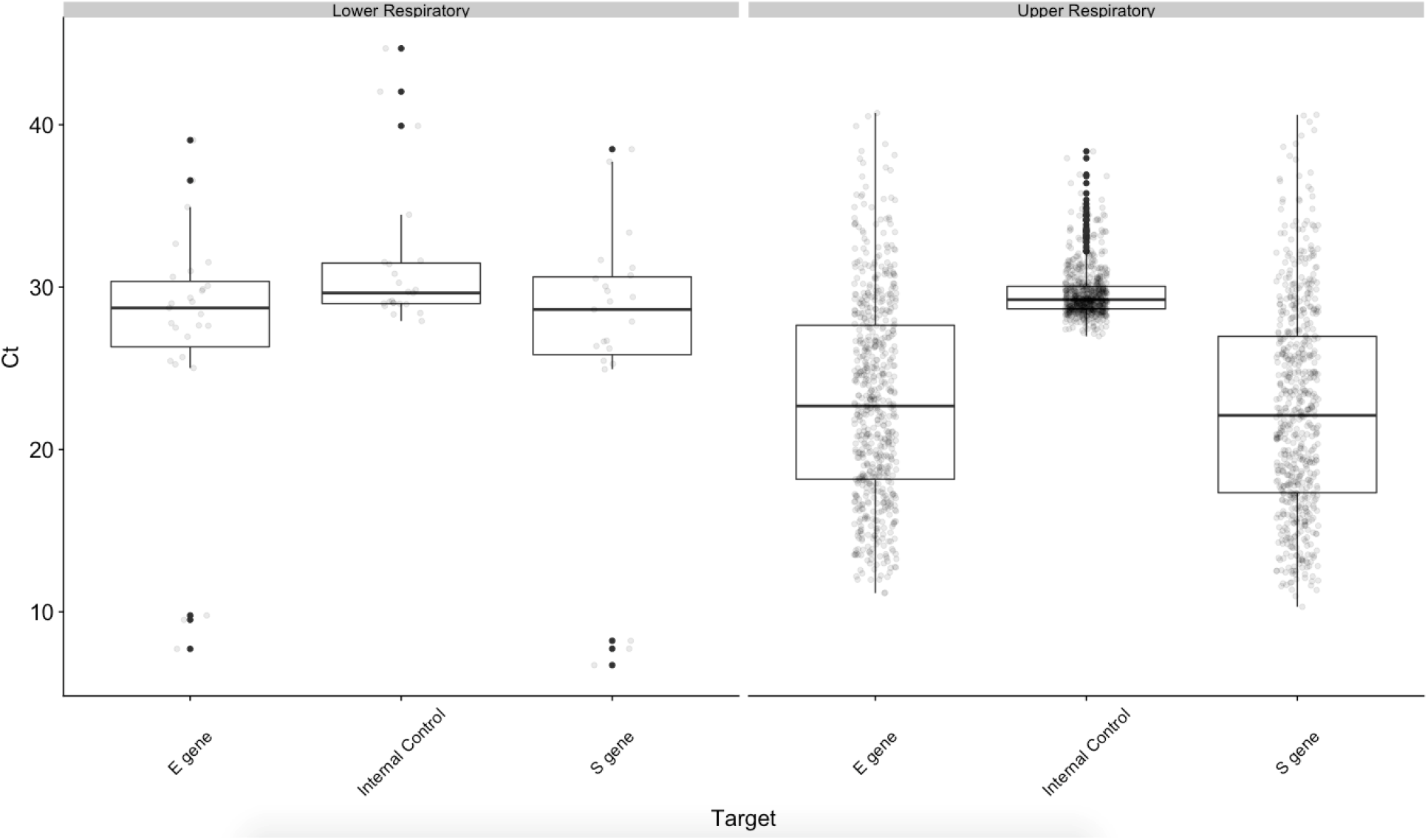
Distribution of Ct values for E gene, S gene, and Internal Control targets for all upper respiratory tract (URT) and lower respiratory tract (LRT) specimens with detected SARS-CoV-2. Mean Ct values between URT and LRT specimens were significantly different for the E gene (p=0.006) and S gene (p=0.03) but not the I.C. (p=0.7), though the much smaller sample size for LRT is noted.

Of 135 patients with repeat testing, only 17 had different results on the second test including 13 patients who first tested negative but subsequently tested positive and three patients who had virus detected in one specimen type but not the other (NP swab but not OP swab detected n=1; NP/OP swab but not NP swab detected n=1, and OP swab but not NP swab detected n=1). One inpatient initially tested positive with B-ßCoV, and then positive with medium viral burden SARS-CoV-2 two days later. Twelve of the thirteen patients who converted from negative to positive were initially tested at the Emergency Department (ED). On repeat testing performed within three days, 5 had low viral burden, 5 had medium viral burden, and 2 had high viral burden on the positive test. Of the ten patients with low-medium viral burden, nine subsequently tested positive as inpatients 1-3 days later, and the tenth tested positive at the ED 3 days later. Two patients tested positive with high viral burden the next day as inpatients. The thirteenth patient tested negative as an inpatient and then with high viral burden as an inpatient seven days later. Of note, while most of the patients in the dataset presented with symptoms and were being tested for suspected infection with SARS-CoV-2, obstetrics and gynecology (OB) patients in the labor and delivery wards were being universally screened for SARS-CoV-2 as a pre-procedural measure to determine if personal protective equipment would be required during interactions with healthcare workers. This group consisted of 102 female patients with a 7% positivity rate.

Means of turn-around-times from test order to result and time-in-lab to result were 19.8 (13.1-26.2) hours and 11.9 (7.0-24.0) hours, respectively. The percentage of tests with detected SARS-CoV-2 increased as the weeks progressed, and settled at approximately 50% from 3/21/2020 to 3/30/2020 (**Figure 4a**). Most of the samples were from the ED (n=911), followed by inpatient wards (n=492) and outpatient clinics (n=113) (**Table 4**), and the highest positivity rate was in the ED with 50% of patients with detected SARS-CoV-2 (p=0.0005). There was a significant difference in the age (p=0.0005) and gender (p=0.005), with lower rates of detected virus in younger patients and female patients. Only 7% of patients 18 years and under had detected virus. Within female patients, older female patients (>55 years, n=346) tested positive with greater frequency than younger female patients (<55 years, n=438) (p=0.001), while this was not the case with male patients in the same age ranges (p=0.09) (**Figure 4b**). This effect was diminished after removing patients from the labor and delivery ward (102 patients, 7% positive) who were being screened universally regardless of symptoms (p=0.03). There was no significant difference in the frequency of positive tests in different race groups (p=0.385).

**Figure 4.**
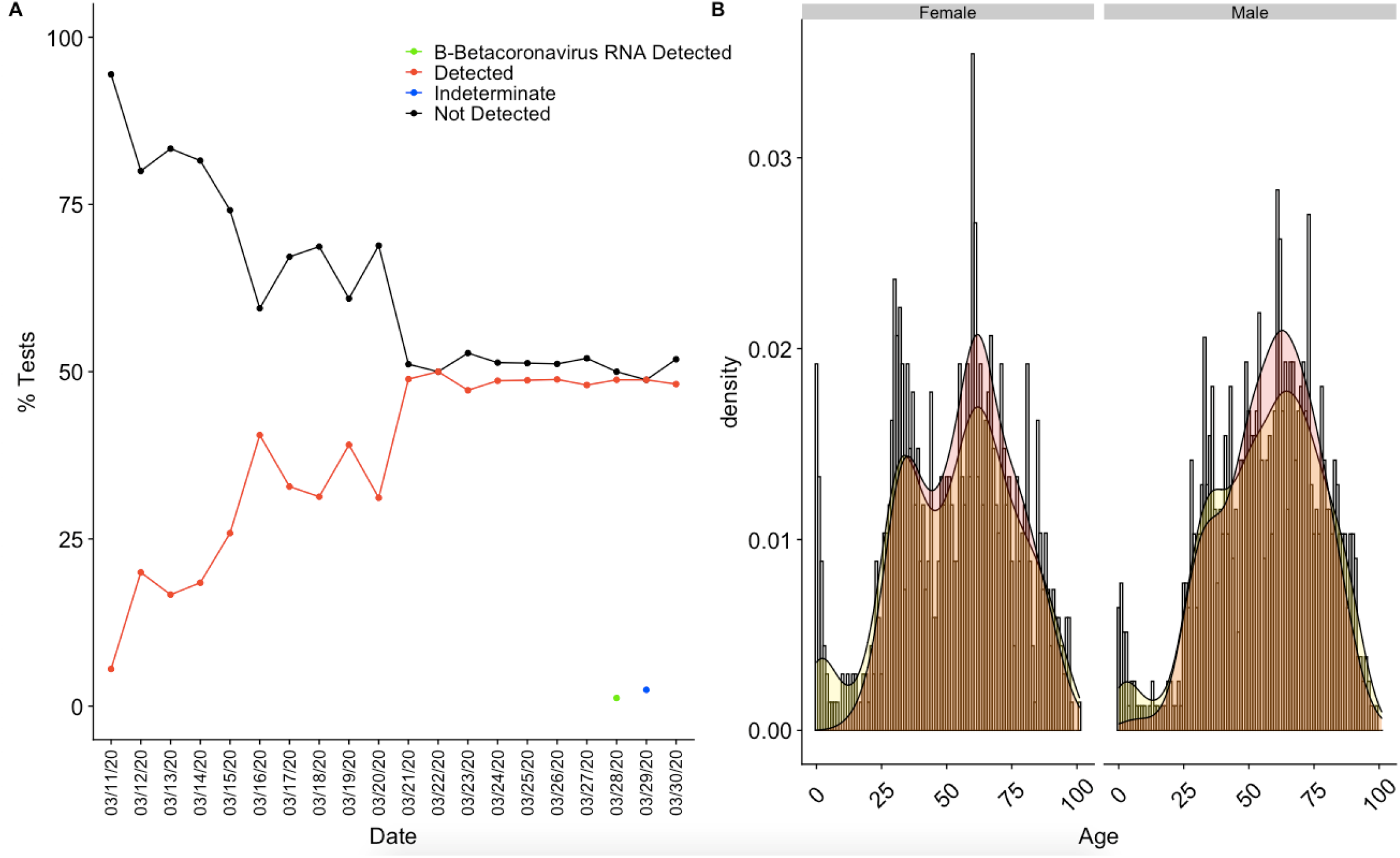
SARS-CoV-2 results by date and distribution by gender and age. **A)** Positivity of URT specimens tested by rRT-PCR at NYP-WCMC over the first three weeks of implementation. **B)** Age distribution histograms with overlays of normalized density curves corresponding to age distribution (yellow) and SARS-CoV-2 positivity (red) in tested patients by gender. Patients that were universally screened at labor and delivery were removed from this analysis.

Lower respiratory tract specimens, including sputum, BAL, and tracheal aspirates, were accepted starting 4/17/2020. As of May 15, 2020, ten sputum, 30 BAL, and 101 tracheal aspirate specimens had been received from 115 patients, with 0%, 13%, and 23%, respectively, showing detectable SARS-CoV-2. Indeterminate results were reported for three tracheal aspirate samples. The mean Ct values for E gene, S gene, and I.C. targets in positive LRT samples (n=27) were 27.3 (7.7-39.1), 26.7 (6.7-38.5), and 31.4 (27.9-44.7), respectively (**Figure 3**). Ct values were not significantly different for the E gene, S gene, and IC targets between positive BAL and tracheal aspirate samples (**Supplementary Figure 1**). The mean number of LRT samples received per day was 7 (range 1-25), which was significantly lower compared to the number of URT specimens tested (mean 85; range 12-176). Given the small sample size, additional statistics on clinical cohort characteristics were not calculated for LRT specimens.

## DISCUSSION

Since the end of December 2019, when China first reported cases of the novel coronavirus disease to the World Health Organization (WHO), SARS-CoV-2 has spread to dozens of countries around the world, including the United States. A rapid and accurate diagnosis of infectious disease is critical for managing outbreaks. Given the increasing number of people infected with SARS-CoV-2 and the lack of any commercially available tests, on February 4, 2020, the FDA in the U.S. opened a pathway that allowed laboratories to implement laboratory-developed tests to meet this diagnostic need. The CDC made their validated kits available through Integrated DNA Technologies, but the kits were very limited in number and were only approved for use with specific instruments, reagents, and controls. Due to a surge in demand for SARS-CoV-2 testing, issues with scaling up numbers of tests per run using the CDC method, and the limited availability of kits, on February 29, 2020, the FDA issued a new policy to help expedite the availability and capacity of diagnostic testing in the U.S. [6]

Altona Diagnostics launched the RealStar® SARS-CoV-2 Real-Time RT-PCR Kit as soon as the disease spread to Europe on February 20. Prototype kits were sent to reference centers for testing and confirmation of functionality with clinical samples [11]. The reagents were designed as a dual target assay and manufactured according to GMP guidelines such that the ready to use kit allowed for rapid detection of all lineage B-betacoronaviruses and SARS-CoV-2 specific RNA in a single reaction. Laboratories could also use the reagents with a wide range of different extraction and real-time thermocycler instruments, allowing for greater flexibility in implementation. The first batch of reagents arrived in our laboratory within five days from ordering on Friday, March 6, 2020, around the same time, we received the WRCEVA RNA reference material. The requirements for obtaining the WRCEVA RNA reference material were 1) that it was used for diagnostics in a CLIA-certified, high complexity lab, and 2) the laboratory was planning to submit an EUA application to the FDA. This RNA reference control was instrumental for the timely development of the EUA test, and notably, many laboratories had difficulties obtaining controls. At the same time, a well-characterized in-house collection of respiratory NP swabs and sputum samples had been collected and saved in the clinical microbiology laboratory, allowing for the preparation of a negative pool of samples for generating the LOD dilution and accuracy samples as described in the Methods section.

Accuracy studies of NP and sputum samples in our laboratory showed excellent overall agreement between the expected and obtained results for contrived clinical specimens and patient samples tested by NYC-DOH. A highly sensitive test is crucial for the detection and identification of SARS-CoV-2 in individuals exhibiting signs and symptoms of a respiratory infection to allow early initiation of therapy. The LODs for the NP and sputum samples were 2.7 gene copies/reaction and 23.0 gene copies/reaction, respectively, suggesting a slightly higher analytical sensitivity for the NP specimens compared to sputum. Overall sensitivity results for this assay by specimen type has been in agreement with the LODs reported in the literature by other studies [11, 12]. Of note, even though the sputum samples had a lower analytical sensitivity, higher viral loads have been reported in sputum specimens compared to NP swab specimens, with LRT samples being the most likely specimen type to test positive for the virus in COVID-19 patients [13]. Based on these favorable validation results, we decided to start routine SARS-CoV-2 testing with the RealStar® SARS-CoV-2 rRT-PCR reagents on March 11, 2020. The entire FDA-EUA validation, followed by a successful go-live testing day, took only four days from receipt of the reagents and RNA reference control in the laboratory.

Overall during this time, we have tested 1,694 URT (40% positive) and 141 LRT (25% positive) specimens, from 1,571 and 115 patients, respectively. The lower number of LRT compared to URT specimens reflects hospital policy, restricting LRT testing to intubated patients that needed clearance of isolation (two negative NP swabs plus one negative LRT specimen) or patients with high suspicion for COVID-19 with repeat negative testing by RT-PCR (two negative NP swabs).

In the cohort of patients tested over three weeks by our assay, positive results were seen more frequently in older males compared to younger and female patients, which has been supported by several studies [14, 15]. Post-menopausal women have been reported to have a greater risk of hospitalization compared to non-menopausal women attributed to the protective effect of estrogen [16]. In this study, older women were more likely to test positive for SARS-CoV-2 compared to younger female patients. However, the difference in detection rate between older and younger women was diminished after removing obstetrics patients screened universally regardless of symptoms, highlighting the importance of restricting comparisons of positivity rates to groups of patients subjected to similar selection criteria and warranting the importance of carefully designed studies.

Among the obstetrics patients, only 7% tested positive for SARS-CoV-2, which is similar to the prevalence (13.5%) obtained for women admitted at delivery at other NYPH campuses [17]. We did not see any differences in the number of positive tests by race, but this was early in the epidemic in New York City. The ED likely had more positive tests since patients tend to be more acutely symptomatic there compared to ambulatory clinics. The percentage of positive tests increased steadily and settled at around 50% three weeks into the epidemic, with later testing on other platforms showing daily positivity rates as high as 75-80% as the epidemic reached its peak in specific boroughs (unpublished data). In this study, 13 patients tested positive after initial negative results in the ED, suggesting they had sufficient symptoms to warrant inpatient admission despite negative testing. This conversion may be due to increased viral burden on subsequent days post-infection, or due to better sampling [18].

In summary, we described the clinical development and implementation of an FDA EUA laboratory validated rRT-PCR test for SARS-CoV-2 in our academic institution, providing a road map to assist others in establishing similar tests. We also described the clinical and testing characteristics of the first cohort of COVID-19 patients admitted to our institution during the early days of the viral outbreak in NYC.

## Data Availability

NA

## ACKNOWLEDGMENTS

We would like to thank all the dedicated medical technologists and health care professionals who performed and assisted in testing at the clinical laboratories of NYPH-WCM. We also thank Dr. Scott C. Weaver, World Reference Center for Emerging Viruses and Arboviruses (WRCEVA), for providing us with viral RNA control material, and Altona Diagnostics for their prompt supply of reagents and support.

## Competing interests

None declared.

## Ethical approval

Obtained.

## Notes

### Competing Interest Statement

The authors have declared no competing interest.

### Clinical Trial

NA

### Funding Statement

No external funding.

### Author Declarations

The IRB Committee at Weill Cornel Medicine (WCM) approved this study.

